# Runners with mid-portion Achilles tendinopathy have greater triceps surae intracortical inhibition than healthy controls

**DOI:** 10.1101/2021.06.25.21259561

**Authors:** Gabriel L. Fernandes, Lucas B. R. Orssatto, Anthony J. Shield, Gabriel S. Trajano

## Abstract

**Objectives:** This study aimed to investigate short-interval intracortical inhibition (SICI) and muscle function in the Triceps surae of runners with mid-portion Achilles tendinopathy (AT).

**Methods:** Runners with (n=11) and without (n=13) AT were recruited. Plantar flexor isometric peak torque and rate of torque development (RTD) were measured using an isokinetic dynamometer. Triceps surae endurance was measured as single leg heel raise (SLHR) to failure test. SICI was assessed using paired-pulse transcranial magnetic stimulation during a sustained contraction at 10% of plantar flexor isometric peak torque.

**Results:** Triceps surae SICI was 14.3% (95%CI: -2.1 to 26.4) higher in AT than control group (57.9%, 95%CI: 36.2, to 79.6; and 43.6% 95%CI:16.2 to 71.1; p=0.032) irrespective of the tested muscle. AT performed 16 (95%CI: 7.9 to 23.3, p<0.001) fewer SLHR repetitions on the symptomatic side compared to controls, and 14 (95%CI: 5.8 to 22.0, p=0.004), fewer SLHR repetitions on the non-symptomatic compared to controls. We found no between-groups differences in isometric peak torque (p=0.971) or RTD (p=0.815).

**Perspective:** Our data suggest greater intracortical inhibition for the Triceps surae muscles for the AT group accompanied by reduced SLHR endurance, without deficits in isometric peak torque or RTD. The increased SICI observed in the AT group could be negatively influencing Triceps surae endurance; thus, rehabilitation aiming to reduce intracortical inhibition should be considered to improve patient outcomes. Furthermore, SLHR is a useful clinical tool to assess plantar flexor function in AT patients.

## 1. Introduction

Achilles tendinopathy (AT) is an overload injury, most commonly seen in the mid-portion of the tendon, which affects both recreational and elite athletes in sports that involve running and jumping^1^. AT is a debilitating multifactorial injury, with high recurrency rates^1,2^. Triceps surae weakness is a key feature in chronic AT and is suggested to be present even before the development of tendinopathy symptoms^3–5^. Muscle weakness has been reported in individuals with AT as reduced maximal concentric and eccentric strength, and endurance. These deficits in muscle strength and endurance are still observed after years of rehabilitation^6,7^, even with improvements in pain and function^5,8,9^. Thus, it is of great clinical relevance to understand the mechanisms underpinning chronic muscle weakness to develop better rehabilitation strategies.

Chronic muscle weakness in individuals with AT could have an origin in the central nervous system. Bilateral sensory and motor deficits (i.e. proprioception and muscle weakness) have been reported in individuals with unilateral tendinopathy, implying involvement of the central nervous system^10–12^. The central nervous system controls the motor action through different inhibitory and excitatory intracortical mechanisms, which will fine tune and adjust the intensity of the stimuli to achieve the required level of force and control^13^. Thus, any changes in cortical mechanisms of inhibition could affect motor performance^13,14^. Short-interval intracortical inhibition (SICI) is a measurement of intra-cortical mechanisms of inhibition, and it is believed to represent post-synaptic inhibition mediated by GABA_A_ receptors^15^. SICI reflects the balance between inhibition and facilitation and can be measured using transcranial magnetic stimulation (TMS), by delivering a pair of pulses to the primary motor cortex. Increased SICI has been reported in patellar tendinopathy patients, suggesting an imbalance within intracortical mechanisms of inhibition; affecting muscle activation and consequently muscle force^14^. Therefore, it is possible that increased intracortical inhibition is an important mechanism affecting muscle force in AT patients.

Although increased SICI has been observed in patellar tendinopathy^14^, it is unclear if this is also present in chronic mid-portion AT patients. Thus, this study aimed to investigate intracortical inhibitory mechanisms in runners with mid-portion AT and triceps surae isometric peak torque, rate of torque development (RTD), and maximal number of single leg heel raises (SLHR) (endurance measure). We hypothesized that runners with chronic mid-portion AT would present with greater levels of cortical inhibition in the triceps surae and lower plantar flexor isometric peak torque, RTD, and number of SLHR than controls.

## 2. Method

### 2.1 Study design and participant selection criteria

This was a cross-sectional study comparing runners with and without mid-portion AT. A sample size of 10 participants was calculated based on the results of Rio et al, 2015^14^ (GPower software parameters: effect size d= 2.63; α err prob: 0.05; power 0.95; n=10). Twenty-four runners were recruited and allocated into two groups: mid-portion AT (n=11) and healthy controls (n=13). Participants’ self-reported height, body mass, age, gender, running mileage, VISA-A, and active motor threshold are reported in Table 1. All volunteers were endurance runners, recruited from local running clubs in the area of Southeast Queensland, Australia.

**Table 1.**
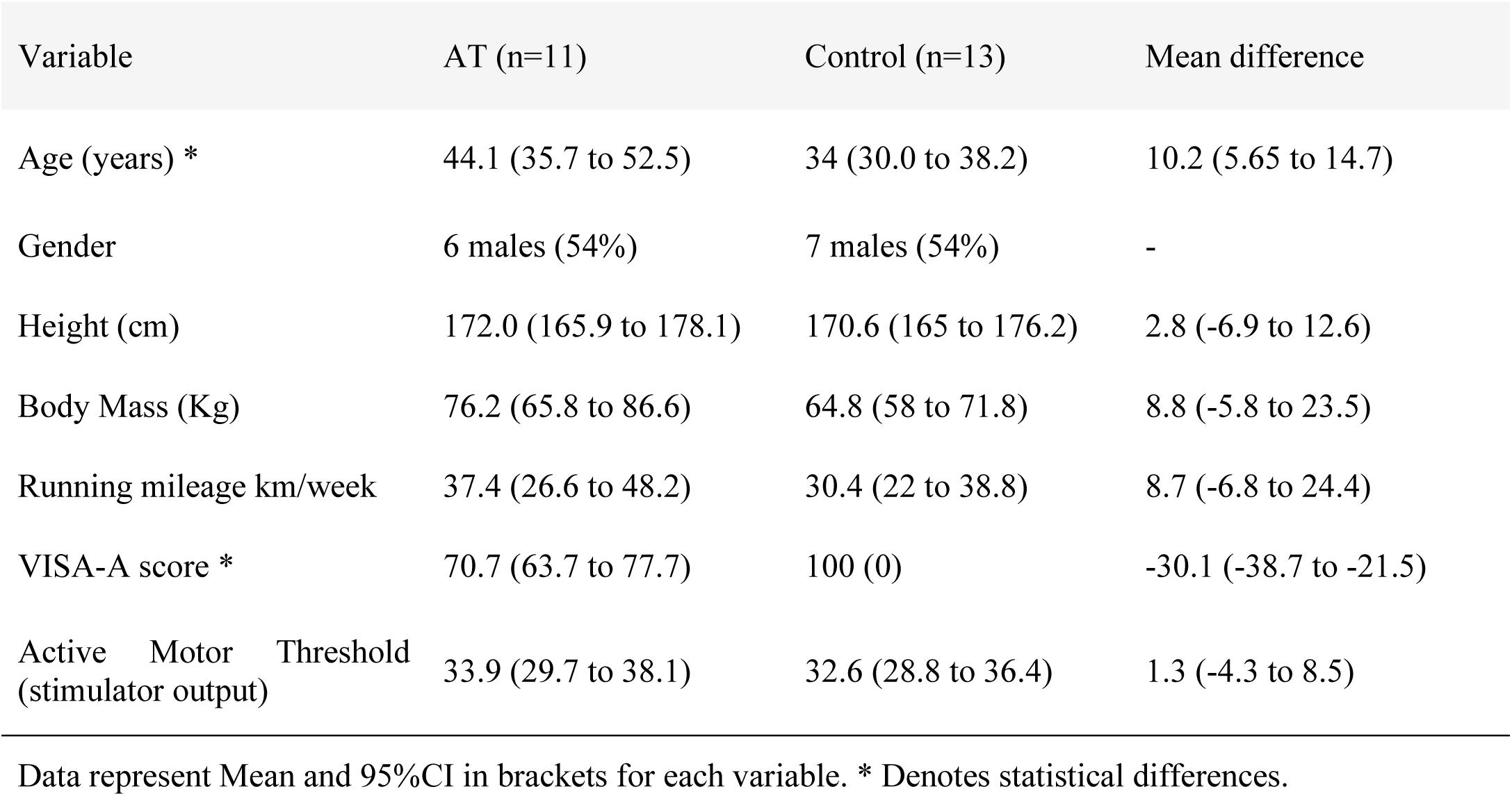
Demographic data for Achilles Tendinopathy and healthy Control group

| Variable | AT (n=11) | Control (n=13) | Mean difference |
| --- | --- | --- | --- |
| Age (years) * | 44.1 (35.7 to 52.5) | 34 (30.0 to 38.2) | 10.2 (5.65 to 14.7) |
| Gender | 6 males (54%) | 7 males (54%) | - |
| Height (cm) | 172.0 (165.9 to 178.1) | 170.6 (165 to 176.2) | 2.8 (-6.9 to 12.6) |
| Body Mass (Kg) | 76.2 (65.8 to 86.6) | 64.8 (58 to 71.8) | 8.8 (-5.8 to 23.5) |
| Running mileage km/week | 37.4 (26.6 to 48.2) | 30.4 (22 to 38.8) | 8.7 (-6.8 to 24.4) |
| VISA-A score * | 70.7 (63.7 to 77.7) | 100 (0) | -30.1 (-38.7 to -21.5) |
| Active Motor Threshold (stimulator output) | 33.9 (29.7 to 38.1) | 32.6 (28.8 to 36.4) | 1.3 (-4.3 to 8.5) |

Data represent Mean and 95%CI in brackets for each variable. \* Denotes statistical differences.

Diagnosis of mid-portion AT was confirmed by an experienced physiotherapist during examination if patients presented with localized mid-portion Achilles tendon pain for more than three months, pain provoked by physical activities in a dose dependent way and had pain with palpation at the mid portion of tendon. Once diagnosis was confirmed, inclusion and exclusion criteria were applied. Inclusion criteria for the AT group were: 1) a diagnosis of mid-portion AT for > 3 months; 2) a running routine of more than twice weekly for > 4 months; and 3) between 18-60 years of age and able to give informed consent. Exclusion criteria for the AT group were: 1) insertional AT; 2) previous rupture or surgery of the Achilles tendon; 3) clinical findings indicating a differential diagnosis for the Achilles tendon pain, such as tendon tear; regular participation in other sports involving high speed running (football, rugby, AFL etc.); 4) VISA-A score > 90 points; and 5) any other musculoskeletal injuries of the lower limb; any neurological disorder; or mental health issues affecting consent. Inclusion criteria for the control group were: 1) to be free of any current musculoskeletal injuries of the lower leg; 2) a running routine of more than twice a weekly for > 4 months; and 3) between 18-60 years of age and able to give informed consent. Exclusion criteria for the control group were: 1) previous injury of the Achilles tendon; 2) regular participation in other sports involving high speed running (football, rugby, AFL etc.); 3) VISA-A score < 100 points; and 4) any neurological disorder; or mental health issues affecting consent. Prior to testing, all participants read and signed a detailed informed consent document and completed the VISA-A questionnaire^16^ and a TMS contra-indication questionnaire. All participants were free of comorbidities such as cardiac, pulmonary, renal, endocrine of gastrointestinal and were not taking any medication for tendon pain or that would affect tendon structure^17^.

This study was approved by the Queensland University of Technology Human Research and Ethics Committee in line with the Declaration of Helsinki. Data collection was conducted during the COVID-19 pandemic and all safety procedures followed local state government policies.

### 2.2 Neuromuscular assessments

#### 2.2.1 Torque measures and single leg heel raise

Plantar flexor isometric peak torque was measured using an isokinetic dynamometer (Biodex Medical Systems, Shirley, New York). For the bilateral AT presentations (n=3), the most symptomatic leg was used and for the control group, the dominant leg was used for testing. Leg dominance was selected by asking the participants what their preferred leg was, and if they were unsure, they were asked which leg they would use to kick a ball. Participants were positioned in a seated position (75 degrees of hip flexion) with their knee straight and with the foot perpendicular to shank. Warm up consisted of 2 × 4 s isometric contractions of each participant’s perceived 20, 40, 60 and 80% maximal voluntary isometric contraction intensity. After warm-up, participants performed at least 3 maximal voluntary isometric contractions, until <5% variation was observed between contractions, and the highest value was used. Participants were instructed to contract as hard and fast as possible and received verbal encouragement and visual feedback of force output during the contractions. Absolute RTD was calculated as the average slope of torque-time curve (0-100 ms) relative to onset of contraction. The onset of contraction was determined by visually inspecting the torque trace and calculated from the time immediately after torque increased from baseline. Relative RTD was calculated by dividing the absolute RTD value by the contraction peak torque. After all neuromuscular and corticospinal assessments were completed, participants were instructed to perform the SLHR endurance test to failure from a flat surface, with each leg. Participants were instructed to lift the heel as high as possible, and return to flat position, as many times as possible^18^. Test was ceased once participants started to perform SLHR test with a reduced range of motion.

#### 2.2.3 Electromyography (EMG)

After skin preparation, bipolar surface electromyography (sEMG) was used to record electrical signals from soleus (SOL), gastrocnemius medialis (GM) and gastrocnemius lateralis (GL), and a reference electrode was placed on the lateral malleolus. sEMG signal was collected during the corticospinal assessments (see below), and amplified (x1000), band-passed filtered (10-1000 Hz), digitized at 4 kHz. sEMG signals were recorded with PowerLab (16/35) and analysed with LabChart 8 software (AD instruments, New Zealand).

#### 2.2.4 Corticospinal assessments

SICI was assessed by measuring motor evoked potential (MEP) amplitudes elicited by TMS over the primary motor cortex. The vertex (Cz) was located according to the 10-20 International System and used as the anatomic reference point for stimulator coil placements. Coil was then, positioned over the motor cortex area related to the lower limb^15,19^. Single and paired pulses were delivered using a double cone coil connected to a Magstim^2^ stimulator (Magstim, Dyfed, UK). Coil was positioned over the motor cortex area, contralateral to the examined limb and oriented to induce a postero-anterior current to the motor cortex. The optimal stimulation site for the triceps surae, was identified using a supra-threshold stimulation intensity with a simultaneous background contraction (10% of peak torque)^20^. The optimal site was considered the area to produce the largest MEP in SOL^15^. Thereafter, active motor threshold was determined using a non-parametric algorithm tool for estimating TMS motor threshold, MTAT2.0^21^. Motor threshold was defined as the minimal intensity of TMS output required to produce a positive MEP in the target muscle in at least 5 of 10 trials. A positive MEP was considered when peak-to-peak amplitude was greater than 0.1mV in SOL^15^.

SICI was investigated using paired pulse stimulation with a conditioning stimulus of 0.8× active motor threshold, followed by a test stimulus of 1.2× active motor threshold, using a 3-ms interstimulus interval^19^. A single pulse was delivered at the same intensity as the test stimulus of 1.2x active motor threshold, as reference to calculate the percentage of SICI. Participants performed a submaximal contraction of 10% of maximal voluntary torque during transcranial stimulation. To ensure correct background torque participants received real-time visual feedback on a monitor positioned in front of them. The decision of whether to initiate the protocol with SICI or single pulse stimulation was randomized for each participant. To ensure MEP reliability across conditions, twenty paired pulses and twenty single pulses were delivered. The results were then compared as an index of intracortical inhibition, whereas the higher the SICI value, the greater the intracortical inhibition. SICI is described as a percentage of the averaged peak-to-peak MEP amplitude and it was calculated using the formula: 100 - (conditioned MEP/unconditioned MEP×100)^19,22^. Testers were not blinded to group during assessment.

### 2.3 Statistical analysis

Repeated measures ANCOVA was used to compare SICI values between muscles of the triceps surae and between groups, as the AT was statistically older than the control group (p<0.01), age was used as covariate in the model. ANCOVA was used to compare between group differences of torque measures, active motor threshold and SLHR using age as covariate to account for statistical difference found in age between groups. Robust paired t test was used to compare SLHR test between legs for participants with unilateral AT (n=8). An alpha level of 5% was adopted for all statistical tests. Hedge’s g was calculated for estimates of effect size (presented as g). Hedge’s g effect size interpretation is as follows: ≤ 0.2 small effect; ≤ 0.5 medium effect; ≤ 0.8 large effect^23^. All statistical analyses were conducted using Jamovi software (Version 1.6, Sydney, Australia). Data is presented as means and 95% confidence intervals and effect size.

## 3. Results

### 3.1 Corticospinal assessments

Triceps surae SICI was 14.3% (95%CI: -2.1 to 26.4; F=5.271, p=0.032, η^2^p=0.201) higher in AT than in controls (57.9%, 95%CI: 36.2 to 79.6; and 43.6%, 95%CI:16.2 to 71.1) (Figure 1); irrespective of the tested muscle (F=0.929, p=0.403, η^2^p=0.042); no muscle × group interaction was observed (F=0.647, p=0.529, η^2^p=0.030) and there was no affect from age in between groups (F=0.991, p=0.331, η^2^p=0.045). Mean active motor threshold did not differ between groups -1.3 stimulator output (−4.3 to 8.5; F=0.197, p=0.661, η^2^p= 0.009).

**Figure 1.**
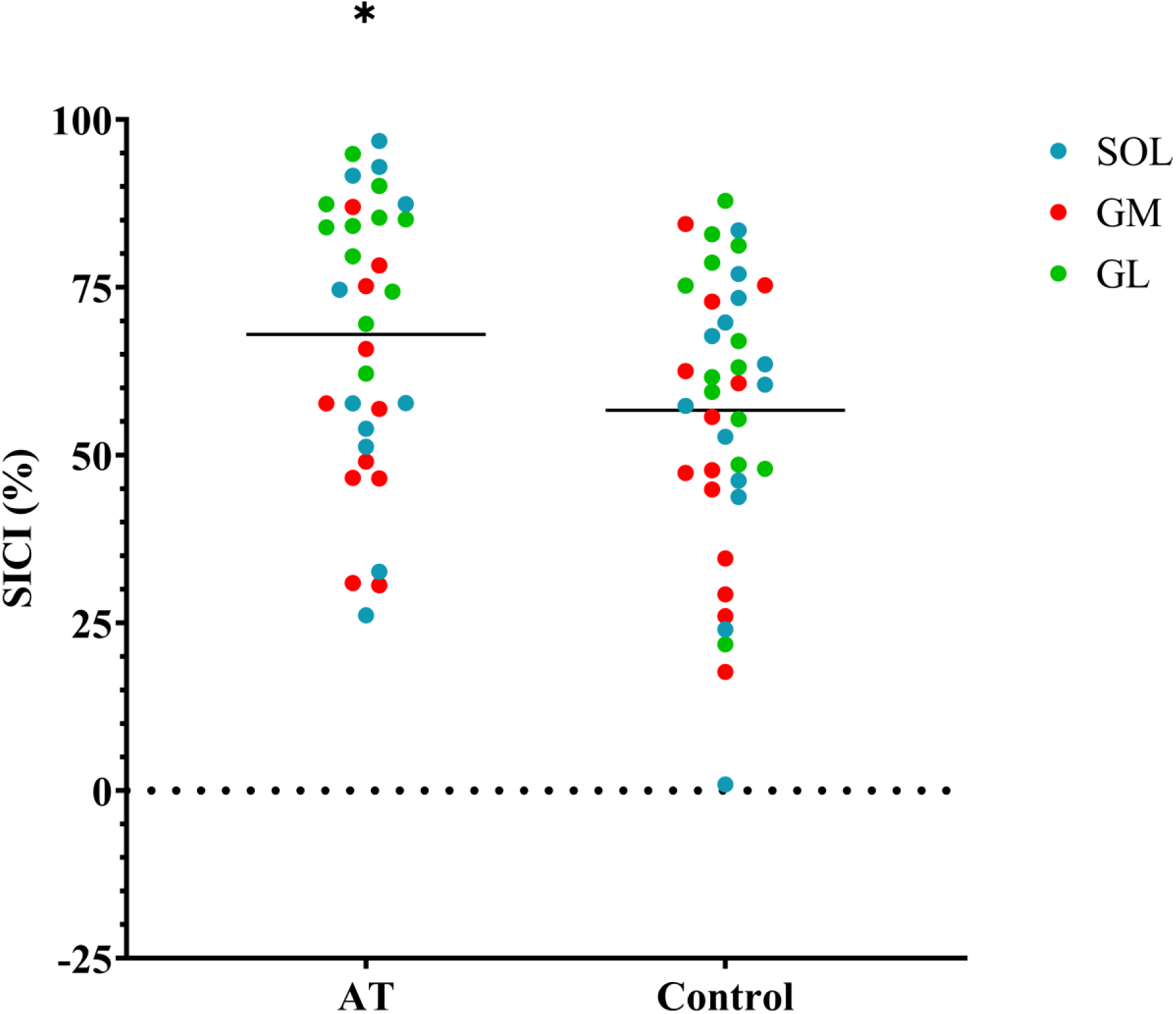
Repeated measures ANCOVA showing SICI individual data for all Triceps surae muscles: Soleus (SOL), Gastrocnemius medialis (GM) and Gastrocnemius lateralis (GL) between groups. The AT group had ∼ 14% more intracortical inhibition than the Control group. SICI was calculated and represents a percentage of inhibition; thus, the higher the SICI value, the greater the cortical inhibition. Data reported as Grand mean. * Denotes statistical difference between groups.

### 3.2 Neuromuscular assessments

We found no differences between groups in any of the torque measures (Table 2); isometric peak torque (F<0.001, p=0.971, η^2^p<0.001); isometric peak torque normalised for body mass (F=1.000, p=0.329, η^2^p=0.045); RTD 0-100ms (F=0.055, p=0.815 η^2^p<0.001); RTD 0-100ms normalised for % peak torque (F=0.150, p=0.702, η^2^p=0.007). For the SLHR test, the AT performed 16 fewer SLHR repetitions on the symptomatic side compared to controls (F=15.64, p<0.001, η^2^p=0.427) and 14 fewer SLHR repetitions in the non-symptomatic side compared to controls (F=11.13, p=0.004, η^2^p=0.382). When analysing side to side differences of the AT, the symptomatic side performed on average, 2.3 less SLHR repetitions compared to the non-symptomatic (g:-0.2, p=0.019), for this analysis, only unilateral presentations were considered. Figure 2 displays data of SLHR values for visual comparison. No participants reported pain during peak isometric torque test or SLHR test.

**Table 2.** Neuromuscular measures between AT and Control

| Variable | AT (n=11) | Control (n=13) | Mean difference |
| --- | --- | --- | --- |
| Isometric peak Torque (N.m) | 110.0 (84.1 to 136) | 111.0 (87.1 to 134) | -0.6 (-35.4 to 34.1) |
| Isometric peak Torque/BM (N.m/BM) | 1.4 (1.1 to 1.8) | 1.7 (1.3 to 2.1) | -0.2 (-0.7 to 0.3) |
| RTD 0-100 (N.m) | 239.0 (151.0 to 326.0) | 224.0 (145.0 to 304.0) | 14.3 (-117.2 to 132.8) |
| RTD 0-100/ peak torque (N.m/ % peak torque) | 2.33(1.5 to 3.1) | 2.12 (1.4 to 2.8) | 0.2 (-0.7 to 1.2) |
| SLHR (Symptomatic vs Control) (repetitions) * | 25.0 (19.3 to 30.7) | 40.6 (35.4 to 45.8) | -15.6 (-23.3 to -7.9) |
| SLHR (Non-symptomatic vs Control) (repetitions) * | 26.3 (19.7 to 32.9) | 40.6 (35.4 to 45.8) | -13.9 (-22 to -5.8) |
| Variable | AT (Symptomatic n=8) | AT (Non-symptomatic n=8) | Mean difference |
| SLHR (AT Symptomatic vs AT Non-symptomatic) (repetitions) * | 26.4 (18.4 to 44.4) | 29.0 (21 to 37) | -2.3 (-3.63 to -1.0) |

Data represent Mean and 95%CI in brackets for each variable.

**Figure 2.**
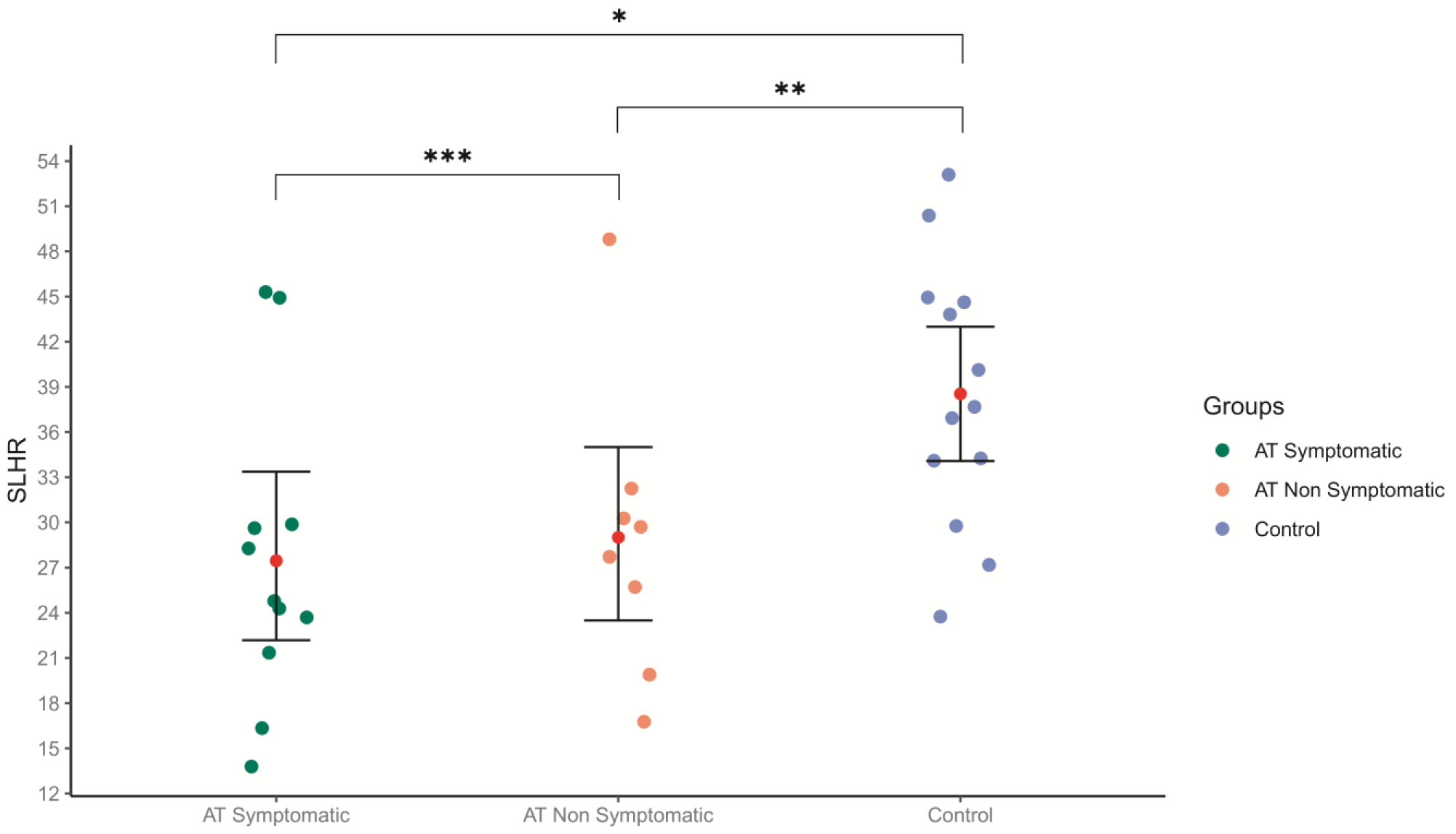
Single leg heel raise (SLHR) endurance test – AT vs Control (individual values). Each point represents individual values of maximal SLHR performed by each participant during endurance test. Both the symptomatic and the non-symptomatic leg of AT group were weaker than the Control group. *AT symptomatic* refers to the most symptomatic leg; *AT Non symptomatic* SLHR used for analysis represent data only from unilateral presentations and for the *Control* group, the dominant leg was used for analysis. Data reported as mean and 95% CI. *; **; *** Denote statistical difference.

## 4. Discussion

This study aimed to determine whether runners with mid-portion AT had: 1) greater intracortical inhibition (measured as SICI) of the triceps surae muscles than healthy controls and 2) lower levels of muscle function (measured as isometric peak torque, rate of torque development and SLHR endurance test). We found that runners with mid-portion AT had higher SICI than controls, accompanied by reduced SLHR endurance but without deficits in isometric peak torque or RTD. The data supports the primary hypothesis that runners with mid-portion AT have greater intracortical inhibition than controls; and partially supports our secondary hypothesis as runners with mid-portion AT have reduced triceps surae endurance. Contrary to the hypothesis, runners with AT were not weaker nor slower in developing torque than controls during isometric contractions.

As expected, SICI was higher in the AT group, outlining increased intracortical inhibition of the triceps surae muscles (SOL, GM, GL), which could be negatively impacting triceps surae endurance. To our knowledge, no other studies have investigated intracortical inhibitory mechanisms, using SICI measures, in chronic mid-portion AT. SICI can be investigated with the target muscle at rest or during a submaximal isometric force (5-10% peak torque); with the latter used in this study, and it measures the activity of intracortical inhibitory circuits within the primary motor cortex (M1) mediated by GABA_A_ receptors,^19^ one of the many circuits that control motor output. Increased cortical inhibition, can reduce the neural drive to the muscle, impacting motor output^24^, such as maximal strength^25^ and endurance^26^. We did not find differences in maximal strength measures; but one study in patellar tendinopathy^14^ reported increased SICI associated with a reduction in maximal voluntary contraction.

They found a reduction of intracortical inhibition, associated with reduction in tendon pain and increased in maximal force following exercise; this study, however, did not have a control group to compare SICI values with. Our results expand these previous findings by demonstrating that patients with mid-portion AT also have higher intracortical inhibition, compared to controls. Additionally, there is evidence^27^ that GL has a 22.8% reduction in activation during submaximal isometric contractions in AT; but our study was not structured to investigate individual muscle differences in neural drive. SICI values can be influenced by age, with an increase in inhibition in older adults^28^, for that reason we used age as a covariate in our statistical model, and still we observed increased SICI in the AT group. This is an important finding to help and better understand the neurophysiological characteristics present in runners with Achilles tendinopathy and its potential impact in chronic triceps surae deficits observed in AT; however, this study cannot address whether increased intracortical inhibition is driving the muscle deficits observed and causing AT, or if this is an adaptation from long-lasting pain and tendon disfunction present in chronic AT. Furthermore, this study does not aim to drive changes in tendinopathy treatment. An interesting point to note, triceps surae heavy resistance rehabilitation programs have yield great improvements in both pain and function in AT patients; heavy resistance training is described in the literature as effective in reducing cortical inhibition^29,30^.

Although there were no differences between groups in measures of isometric peak torque, the AT group showed a reduced capacity during the SLHR endurance test, suggesting a deficit in triceps surae endurance. The lack of group difference in plantar flexor isometric peak torque is in line with the result of other studies^27,31^. In contrast, weakness in plantar flexor dynamic peak torque has been widely reported in AT^5,9,12^. Perhaps, as observed in other chronic musculoskeletal disorders, such as chronic hamstring strains^32^, chronic AT presents deficits in dynamic but not in isometric peak torque. It is interesting to note that the AT group was on average 8.8 kg heavier that the control group, and yet there were no differences in torque measures in absolute or in normalised values. To the authors’ knowledge, studies have not yet compared concentric, eccentric, and isometric torque measures from the same group of AT patients, against a control group. Further research analysing different contraction types is necessary to fully characterize triceps surae`s deficits in AT and help guide better rehabilitation strategies.

Explosive plantar flexor torque, measured as the RTD, was not different between groups in absolute or normalized values, suggesting chronic AT has no impact on this important parameter. A recent meta-analysis^9^ found conflicting evidence about reductions in RTD in AT; perhaps other factors could be impacting hop distance, rather than explosive force, such as increased tendon compliance observed in chronic AT^31^. Our study compared RTD from the most symptomatic leg of AT against healthy controls, whereas all other studies investigating RTD in AT, compared RTD between symptomatic and non-symptomatic legs of the same subjects. As a consequence, there is limited evidence to compare our data to.

Kinesiophobia (fear of movement) influences endurance recovery in AT patients; patients with higher levels of kinesiophobia recovered less of their muscle endurance in a 5-year follow-up^2^. This interesting finding shows how fear of moving in a certain way or using a specific body part can impact the neural drive to the muscle, possibly involving inhibitory mechanisms within the cortex, as observed in the AT group.

The SLHR is a common clinical test of triceps surae endurance^18,33^ and function in AT patients^33^. Our data showed a reduction of 38% in triceps surae endurance on the symptomatic side and 34% on the non-symptomatic side in unilateral presentations, compared to controls. Although we found a difference between sides of the AT group, the effect size was only small. Participants performed SLHR first on their symptomatic/dominant leg, followed by their non-symptomatic/non-dominant leg. The non-symptomatic leg of the AT group was performed immediately after the symptomatic leg. Thus, it is possible that the difference observed between the symptomatic and the non-symptomatic observed in the AT group were attenuated by mechanisms of central fatigue.

Studies that have adopted the SLHR endurance test^2^ or calculated the total work done in dynamometry^5^ for endurance measurement have also observed a reduction in endurance capacity compared to the non-symptomatic side and to controls, respectively. Bilateral deficits in unilateral presentations have been widely described in chronic AT^9,12^. These deficits on both symptomatic and non-symptomatic sides, emphasize that the non-symptomatic leg should not be used as comparison for neuromuscular function in AT. It is possible that the degree of cortical inhibition present in our AT participants, is insufficient to impact muscle performance during a brief maximal voluntary isometric contraction, but sufficient to reduce performance during the SLHR test, once additional inhibitory impacts (sensory inhibitory drive to motor neurons or higher centres) are added. Furthermore, it would also be relevant to investigate the impact neural mechanisms would have in muscular responses to fatigue in AT.

Our AT group had greater running mileage/week than controls (Table 1). AT is an overloading injury and higher mileage could be a contributing factor for the development of AT. It is suggested that, amongst other factors, load magnitude and recovery time, may influence tendon response and help develop tendinopathy^34^.

The literature shows no association between tendon structure and levels of tendon pain and dysfunction^35^; however, not having scanned participants tendons could be considered a limitation of this study. In this study we had 3 runners with bilateral AT and 8 runners with unilateral AT. It is still unknown if runners with bilateral AT present specific cortical changes in comparison with runners with unilateral AT. Futures studies investigating neurophysiological differences between unilateral and bilateral AT could help to fill this gap.

Unilateral AT presentations displayed bilateral deficits in muscle endurance; yet, without presenting any symptoms; thus, having scanned participants bilateral tendons, would have helped profiling our participants for further analysis and understanding. Testers were not blinded for group, which is also a limitation of this study.

## 5. Perspective

Runners with mid-portion AT have greater intracortical inhibition to the triceps surae muscles accompanied by a bilateral reduction in triceps surae endurance, despite no deficits in isometric peak torque or explosive torque. The increased SICI observed in the AT group may be negatively influencing triceps surae endurance; thus, rehabilitation strategies aiming to reduce intracortical inhibition should be considered for better outcomes. Furthermore, the SLHR test, is a useful clinical tool to assess triceps surae function in patients with mid-portion AT as an outcome measure of improvement.

## Data Availability

Data material will be send as requested

## Acknowledgments

The authors declare not having received any funding for this study.

The authors thank the following for their contribution to the development and achievement of this research:

Miss Charmaine Enculescu who assisted with figure plotting;

All volunteers who participated in this study.

